# Explainable-AI to Discover Associated Genes for Classifying Hepato-cellular Carcinoma from High-dimensional Data

**DOI:** 10.1101/2022.08.14.22278747

**Authors:** Easin Hasan, Fahad Mostafa, Md S Hossain, Jonathon Loftin

## Abstract

Knowledge-based interpretations are essential for understanding the omic data set because of its nature, such as high dimension and hidden biological information in genes. When analyzing gene expression data with many genes and few samples, the main problem is to separate disease-related information from a vast quantity of redundant data and noise. This paper uses a reliable framework to determine important genes for discovering Hepato-cellular Carcinoma (HCC) from micro-array analysis and eliminating redundant and unnecessary genes through gene selection. Several machine learning models were applied to find significant predictors responsible for HCC. As we know, classification algorithms such as Random Forest, Naive Bayes classifier, or a k-Nearest Neighbor classifier can help us to classify HCC from responsible genes. Random Forests shows 96.53% accuracy with *p <* 0.00001, which is better than other discussed Machine Learning(ML) approaches. Each gene is not responsible for a particular patient. Since ML approaches are like black boxes and people/practitioners do not rely on them sometimes. Artificial Intelligence(AI) technologies with high optimization interoperability shed light on what is happening inside these systems and aid in the detection of potential problems; including causality, information leakage, model bias, and robustness when determining responsible genes for a specific patient with a high probability score of almost 0.99, from one of the samples mentioned in this study.

## I. Introduction

Cancer deaths from hepatocellular carcinoma (HCC) are the most common cause of mortality around the globe. HCC causes more than 85 percent of all primary liver cancers. It is the sixth most prevalent cancer worldwide and the second leading factor in cancer-related fatalities [1]. The high mortality rate of HCC results from metastasis or the development of newly generated tumors within the sick liver. Research has revealed that 90% of all cancer-related fatalities are caused by metastasis. Since HCC’s genomic changes and gene expression patterns are so diverse among patients, it is nearly impossible to pinpoint the mechanisms and pathways of the disease. That is why ineffective therapy is to blame for poor patient outcomes in the HCC patient group. Recurrence after surgery is a major factor in HCC’s poor prognosis, and there are currently few therapeutic approaches that successfully reduce recurrence due to metastasis. As a result of this difficulty in identifying HCC patient subgroups at high risk of developing metastatic illness in advance, one of the key reasons for this study. The efficacy of a metastatic signature made up of 153 genes that might identify HCC patients as a risk classifier for HCC recurrence and survival was examined using two independent cohorts totaling 386 HCC patients [2]. But this study wasn’t enough to predict HCC in the early stage of cancer development which motivated us to predict the genes responsible for HCC so that doctors and health practitioners can take the necessary steps to prevent HCC in the early stage. So, if they can identify the responsible genes for HCC existing in the patient’s body, many lives can be saved from this cancer disease. According to the conventional tumor evolution model, a primary tumor is initially benign but develops mutations over time, allowing a small number of tumor cells to spread. To enhance patient survival, early diagnosis of tumors that have already mutated is extremely important. Using genes whose copy counts correspond with gene expression and cancer development as the standard for HCC driver genes, Roessler et al. employed an integrated strategy to find these genes, even though the mechanisms underlying the aggressive cancer HCC genesis and progression are poorly understood [3]. This study directed us to apply state-of-the-art Artificial intelligence techniques to determine the responsible genes for developing HCC. So from a patient’s genomic information, if health practitioners can identify those responsible genes, individuals can begin medical therapy when the disease is still in its early stages. Although surgical resection and liver transplantation are possible, the recurrence rate is significant, and surgery is usually out of the question since the disease has progressed too far. Given that patients with HCC, especially those incompetent for surgical resection or liver transplant, it is crucial to identify key drivers and potential treatment targets [4]. Zhao et al. show the viability and strength of a novel approach by identifying key pathways associated with prognostically significant HCC subtypes using well-defined patient samples and integrated genomics [3]. But they only used unsupervised clustering algorithms to find the key pathways, which can be improved significantly by applying the machine learning algorithms to the high dimensional genomics data to figure out the key genes which cause the HCC disease. It is common practice to extract a biological sample and then use microarrays to express the genes. Statistical approaches are used to assess the data and discover meaningful meaning that biologists can utilize to give them biological significance once the data have been transformed into numbers. Numerous genome-wide tools, including microarrays and, more recently, next-generation sequencing platforms, have been used to analyze thousands of clinical HCC samples in an effort to identify promising treatment targets [4]. But, there are minimal applications of machine learning methods in genomic datasets for identifying the key genes besides the bioinformatic analysis. Wang et al. discovered 13 clinically significant target genes with therapeutic promise utilizing genome-wide growth depletion screens and combining real HCC tumors’ expression data and clustered regularly spaced short palindromic repeats [5]. So. there is much possibility to discover the significant target genes by applying machine learning algorithms in addition to the bioinformatics analysis. Lu et al. investigated Numb mRNA expression in tumors and surrounding healthy tissues using either 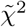 or Mann-Whitney U-tests on a microarray dataset of 241 HCC patients to determine the relationship between clinicopathological traits and HCC subtypes [6]. But machine learning methods could be very useful for analyzing a high-dimensional dataset to determine additional marker genes which cause HCC. Identifying genes that could serve as biomarkers is critical to diagnose patients better early on and enhance the effectiveness of treatment options while also increasing the likelihood of a positive outcome. A microarray is a kind of experimental setup where probes are created or attached to solid substrates to expose them to the target molecules. The microarray data set holds experimental data in a matrix, where columns typically correspond to sample IDs and rows typically to the names of genes or probes. Microarray genomic data analysis is very useful for finding the differentially expressed long non-coding RNA (lncRNA). According to Chen et al. bioinformatic analysis in the liver tissue in the brain-dead donor liver transplantation, microarray probes effectively identified hundreds of transcripts by displaying differentially expressed lncRNA and circRNA profiles [7]. So, using machine learning techniques, we may decipher nuanced connections and identify intricate patterns in microarray data. Healthcare researchers utilize the study of microarrays and gene expression as a tool to learn about diseases and how to remedy them. It may provide important information about highly complicated diseases like HCC. Due to its poor prognosis and ineffective response to systemic therapy, HCC ranked fourth among the top causes of cancer-related fatalities in 2018. Considering this situation, the HCC survival rate is still below tolerable levels. As a result, it is essential to keep looking for possible HCC prognostic and therapeutic targets by improving our comprehension of its cellular biological processes [8]. The genetic machinery responsible for metastasis may be hard-wired into tumors from the start, which motivates tumor profiling to forecast patient progress. Benefits of Machine Learning in Health Care.

Machine learning techniques have lately become more precise and effective than conventional parametric algorithms when used for large area modeling and working with high dimensional and complex data sets [9], [10], [11]. Since these algorithms don’t rely on data distribution assumptions like normality, they are more accurate, efficient, and effective [12] [13]. However, because of the numerous parameters that need to be modified and the complexity of implementing them, some machine learning systems (such as neural networks and support vector machines) are sophisticated [14], [15]. Furthermore, these algorithms frequently overfit the data [16]. For two class situations, new work shows that random forests are equal to a kernel operating on the true margin in distribution space [17]. The symmetry of the kernel is said to be enforced by randomness (poor correlation), while strength increases a desirable skewness at abruptly curved boundaries. This should clarify the dual function of correlation and strength. Understanding may also be aided by [18] theoretical framework for stochastic discrimination. The random forest classifier has now been tested against the probabilistic and k-NN classifiers. It is based on probability models with strong assumptions of independence. Probability models may be created using Bayes’ theorem. Despite their simple design and assumptions, naive Bayes classifiers [19] optimality have performed excellently in various challenging real-world settings. However, a comprehensive comparison with different classification algorithms in 2006 revealed that more advanced methods, such as boosted trees or random forests, outperformed Bayes classification. The Naive Bayes Classifier [20] model was employed to determine if it yielded the same conclusions as the Gene Expression data. The random forest method does not require the researcher to propose any particular model structure. This is essential in early genome-wide or candidate area prospective studies where the feature’s genetic structure is ambiguous. However, combining a vast number of genes and relatively small microarrays introduces new challenges for statistical models. Furthermore, they are becoming increasingly popular due to their ability to tackle large numbers of genes without conventional feature selection, robustness to outliers, and extensive application in microarray data analysis. Santos et al. employed logistic regression and neural network classifiers to classify the 165 patients in the Coimbra Hospital and University Centre database, with NN achieving an accuracy of 75.2% and LR of 73.3%, respectively [21]. The proper preprocessing of the dataset might considerably enhance their model to achieve better accuracy, inspiring us to use the cutting-edge data reduction technique PCA in our HCC dataset. Acharya et al. introduced a hybrid system that used three algorithms, including linear discriminant analysis to minimize the number of features, a Support Vector Machine for classification, and a Genetic Algorithm to improve the model, and produced an accuracy of 90.30%, specificity of 96.07%, and sensitivity of 82.25% [22]. Models must be understandable by users if humans are to trust AI technologies. AI interoperability sheds light on what is happening inside these systems and aids in the detection of potential problems, including causality, information leakage, model bias, and robustness. The concept comes from a work published in 2016 [23] in which the researchers perturb the initial data points, feed them into the black box model, and watch what happens. The approach then adjusts the weights of those additional data points based on how close they are to the original point. It uses those sample weights to fit a substitute model, such as linear regression, to the data set with variations. The newly trained explanation model can then be used to explain each original data point. Local interpretable model-agnostic explanations(LIME) provide a general framework to decipher black boxes and explain the” why” behind forecasts or suggestions made by AI. Local Interpretable Model-Agnostic Explanations is known as LIME which was studied by many researchers for improving diagnoses of patients [24], [25], [26]. In this study, we are interested in how doctors and patients can trust machine learning prediction when each patient is different from the other, and multiple parameters can decide between Hepato Cellular Carcinoma or not. To solve this problem, LIME has been used in the test model. The method of explanation should be applicable to all ML models. The researchers term this as the explanation being model-agnostic along with the individual predictions, and the model should be explainable in its entirety, i.e., a global perspective was considered.

In this research, a unique framework is established for gene expression data of HCC to figure out the genes responsible for HCC. The accuracy of ML models is determined where the best model, among several classification algorithms, and the trained model pass thru an AI-explainable technique. In brief, we want to find patterns in the gene changes to assess whether they are normal or indicative of a disease and identify the circumstances under which a gene transforms from a healthy state to a pathogenic state using several machine learning models. This article organizes as follows: section II describes the framework for this study with a brief discussion of the mathematical methodology of Random Forest, logistic regression, Naive Bayes classifier, and k-nearest neighborhood classifier models. To select the important variables, Gini Index and entropy with information are used. To visualize and reduce dimensionality, a heat-map and principal components are used extensively. After using various classification techniques to predict HCC, ROC and bio-statistical analyses are reported. The local interpretable AI method, LIME, was used to interpret the responsible genes for a particular HCC patient. Section III is dedicated to the results and discussion of ML classifications and AI explanations. Finally, section IV concludes this study.

## II. Methodology and Framework

### A. Samples from Gene Data

Data was collected from the national center for biotechnology information. The title of the data is Gene expression data of HCC with ID GSE14520 [27], and the access date of this data set was March 01, 2022. To study the gene expression patterns in HCC patient tumor and non-tumor tissue matched with healthy donor liver tissue, Affymetrix microarray profiling was used by lab technicians. Using a single channel array technology, tumors and matching non-tumor tissues were assessed independently for gene expression profiling. On Affymetrix GeneChip HG-U133A 2.0 arrays, data providers used the manufacturer’s instructions to evaluate tumor and non-tumor samples from 22 patients in cohort 1, as well as normal liver samples. They measured the fluorescence intensities by using GCOS Affymetrix software and an Affymetrix GeneChip Scanner 3000. On the 96 HT HG-U133A microarray platform, all samples from cohort 2 and 42 tumors and paired non-tumor samples from cohort 2 were processed. So, we extracted the gene expression microarray data GSE14520 from the NCBI GEO website. Four hundred forty-five samples were taken from patients between 2002 and 2003 at the Liver Cancer Institute (LCI), Fudan University in China, and the Liver Tissue Cell Distribution System (LTCDS), the University of Minnesota in the United States. The dataset contained 222 cases and 212 controls; however, no case-control data were provided for 11 cases. The majority of HCC patients had a history of hepatitis B infection (96.31%).

### B. Mathematical Framework Machine Learning Approaches

Let us consider the predictor gene data matrix is **X** ∈ **R**^**nxd**^ and target/response variable is **T** ∈ **R**^**d**^. The prepossessed micro-array data set was retrieved from the NCBI GEO repository and cleaned using the R software’s limma package and Bio conductor. The dimension of this dataset is 445 by 22268 with 445 samples and 22268 genes. Additionally, of the data set’s 46 phenotypic features, tumor and non-tumor tissue types are employed as response variables and are factorized into 0 and 1 to facilitate the analysis. The data set was additionally cleaned to eliminate any NA, and a response variable column was added to the gene dataset along with sample ID matching between the phenotypic dataset and the gene dataset.

This data set was used for machine learning and statistical analysis to predict Hepato Cellular Carcinoma. Four different classification methods were used such as random forest, naive bayes classifier, k-NN and Logistic Regression, and the final results in the form of plots such as Confusion Matrix, ROC analysis were compared to get the results.

Data mining is a relatively efficient way of analyzing data that is used to discover patterns and connections in gene data that may suggest genuine interference. A newly refined one-size-fits-all technique will be effectively used in microarray data prediction. The decision tree is the most well-known and practical approach for discovering information from massive and complicated data sets. Moreover, it is straightforward to understand and implement.

#### 1) Variable selection for classification and statistical analysis

To identify important variables/genes, Principle Component Analysis [28] was employed, and it was compared to the Variable Importance ranking [29]. Since RF is used for classification, and RF is the average of multiple decision trees. As a result, the following discussions are incorporated into the framework.The data sets must first be divided into parts in order to form an intermediate decision tree with roots nodes at the top, which is fundamental to decision trees. Then, the decision tree’s stratification model leads to the final result through the tree pass-over nodes. A comprehensive discussion of Entropy, Gini Index, and Information Gain, as well as their roles in the Decision Trees method implementation, may be found here ([30], [31]). Furthermore, because many factors influence decision-making, the significance and impact of each factor must be studied. The root node is assigned the required feature, and the node division is traversed downwards. At each node, descending downwards reduces impurity and uncertainty levels, resulting in enhanced classification or an exclusive split. Splitting measures such as Entropy, Information Gain, and the Gini Index have been used to solve the problem. Entropy quantifies the impurity or randomness of data points. It is a value between 0 and 1. Entropy close to 1 indicates that the model has a higher level of disorder. The concept of entropy is crucial for computing Information Gain. By determining which feature provides the most information about the classification based on the idea of entropy, Information Gain is used to determine which feature provides the most information about the classification, with the goal of reducing the amount of entropy starting from the top (root node) to the bottom (leaves nodes). Mathematically, Entropy is defined as follows:

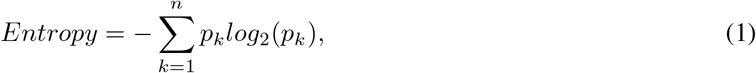

where *p*_*k*_ denotes the probability that it is a function of entropy. The Gini Index is defined as follows: The Gini index ranges from 0 to 1. A Gini Index of 0 corresponds to absolute classification, meaning that all of the items belong to a single class or that only one class is present. A Gini Index of 1 shows the uneven distribution of components across several categories. A Gini Index of 0.5 indicates that items in some classes are distributed equally. The Gini Index can be represented mathematically as:

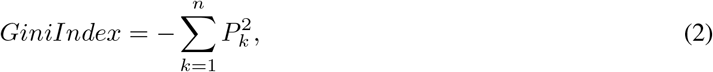

where *P*_*k*_ is known as the chance that an element will be assigned to a specific class. Each feature’s importance is determined by the Gini Importance or Mean Decrease in Impurity (MDI) method, which adds up all of the splits (across all trees) that include the feature in proportion to the amount of samples it splits. This idea was used in the RF algorithm after discussing the other classification algorithms below.

#### 2) Classification of Gene Expression Data

To classify HCC from gene expression data, four different Machine Learning techniques are applied. The foremost technique is Naive Bayes Classifier, which is asymptotically equivalent to Logistic Regression if the Naive Bayes assumption holds. Another well known technique is k-Nearest Neighbor(kNN). The Euclidean distance between the test samples and the training samples is used as the basis for kNN’s determination of the class level for the test samples. As a result of the microarray profiling, gene expression patterns in healthy donor livers, as well as tumor and paired non-tumor tissue from HCC patients, were analyzed. kNN classifies whether it is tumor and paired non-tumor tissue of HCC patients or not based on the Euclidean distance. Finally, Random Forest method is applied and its result produces from the aggregation of the decision trees.

### 2.1. Naive Bayes classifier for HCC Classification

Although the Naive Bayes classifier is not linear in general, it corresponds to a linear classifier in a given feature space provided the probability likelihood factors *p*(*t*_*i*_|*c*) come from exponential families. The data in this case comes from HCC, and the label is either cancer or not. Whether or not we are aware that a patient has cancer, the Naive Bayes assumption states that the variables in the data are conditionally independent, both liver cancer and non-liver cancer are drawn independently at random. Although this assumption can be fragmented, the resulting classifiers can still perform well in real-world applications [32], [33]. Let’s assume the Naive Bayes assumption holds for now and define the Bayes Classifier by:

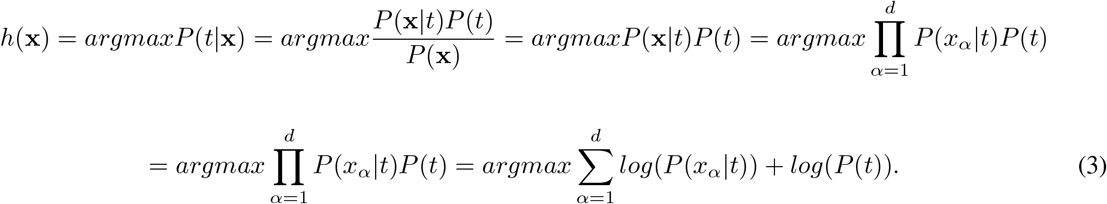

One dimension estimating *log*(*P* (*x*_*α*_|*t*)) should be considered, because it is easy to calculate. Now suppose that *t*_*i*_ ∈ (−1, +1) and features are multinomial. The goal is to show that,

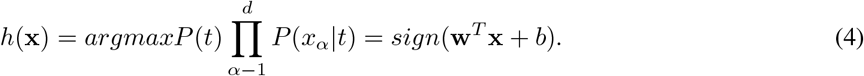

This is

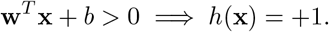

Since, 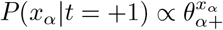 and *P*(*T* = +1) = *π*_+_:

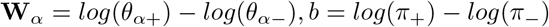

One can compute the **w**^*T*^ **x** + *b* by applying the above for performing classification. By further simplification,

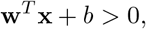

which implies

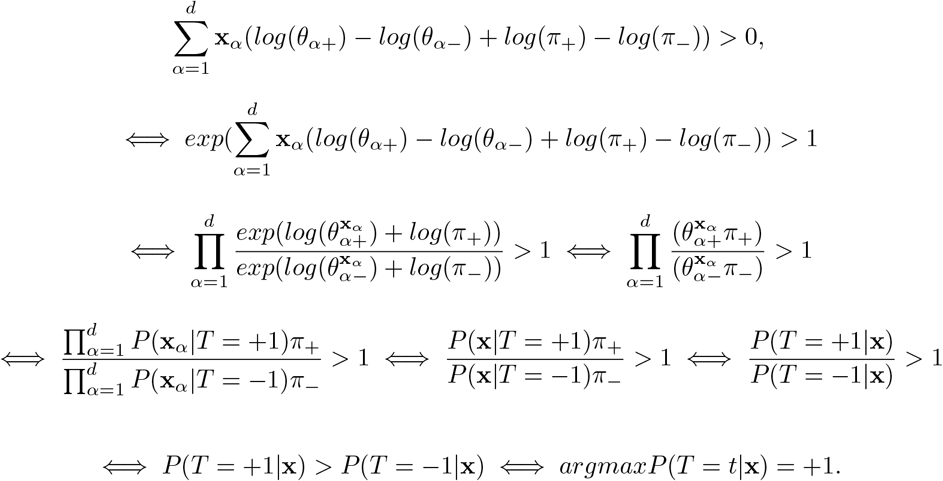

Iff Naive Bayes predicts +1 this demonstrates that point **x** is located on the positive side of the hyperplane. In this gene expression data set all the variables are continuous, therefore Gaussian Naive Bayes can be considered. Thus, Naive Bayes Classifier and Logistic Regression produce asymptotically identical models, if the Naive Bayes hypothesis holds.

### 2.2. Logistic Regression classifier for HCC Classification

The Logistic Regression model [3, 4] is used to model the relationship between a binary target variable and a set of independent variables. In logistic regression, the model predicts the Logistic Regression transformation of the probability of the event, which is used for the high dimensional gene expression data [34]. The following mathematical formula is used to generate the final output:

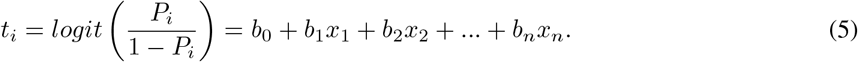

The odds ratio is represented by *P*_*i*_ in the equation above, and its formula is as follows:

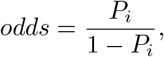

where, *P*_*i*_ stands for probability of “presence of Hepato Cellular Carcinoma”, and 1 − *P*_*i*_ is for “absence of Hepato Cellular Carcinoma”. As a result, the predicted values from the above model, the log odds of the event, can be transformed into the probability of an event as follows:

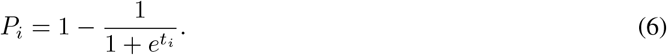

In matrix-vector form, *t*_*i*_ = *b*_0_ + **b**^*T*^ **X**. Then (*b*_0_, *b*_1_, …, *b*_*n*_) is fitted thru Maximum Likelihood Estimator. Then the log-likelihood function is 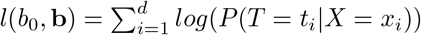. Therefore,

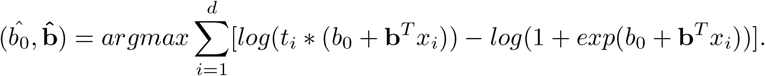

Another well known classifier is the k-Nearest Neighbor classifier which depends on a distance metric [35], [36]. The more accurately the metric captures label similarity, the more accurate the classification is determined. k-NN is discussed in the next sub section.

### 2.3. k-NN classifier for HCC Classification

k-NN is based on a distance metric. The Minkowski distance is the most popular option. Mathematically the Minkowski distance is denoted by *d*(**x, t**) for data point **x** and **t**, and defined by

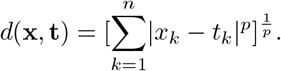

When *p* = 2, it becomes the *l*_2_ distance. Here, **t** are the test points, and the set of the *k* nearest neighbors of **t** is given as *S*_*t*_. Formally *S*_*t*_ is defined as a subset of the data set *D* s.t. |*S*_*t*_|= *k* and (**t**^′^, **x**^′^) ∈ *D* − *S*_*t*_, therefore the metric has the following property:

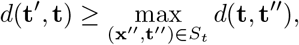

i.e., every point in *D*, but not in *S*_*t*_, is at least as far away from **t** as the furthest point in *S*_*t*_. We can then define the classifier *c*(.) as a function returning the most common label in *S*_*t*_. Specifically,

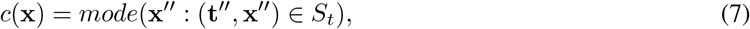

where mode(.) means selecting the highest occurrence label. Here *k* is determined before training the algorithm, and a good solution is to return the result of k-NN with smaller *k* [37]. Moreover, for binary classification, another method known as the random forest is based on the average of many decision trees. Each tree is weaker than the combination of all trees, but together they are powerful. Therefore, it yields better performance.

### 2.4. Random Forest classifier for HCC Classification

Random Forest is a supervised learning approach that employs a tree-based ensemble in which each tree is dependent on a set of random variables. It is a combination of the two classification trees that differ in two important respects. Each tree is first fitted to a random bootstrap sample selected from the entire dataset. To obtain the bootstrap sample, we randomly sample microarrays from the underlying data with replacement until our sample is the same size as the original. Out-of-bag data refers to microarrays that did not make it into the bootstrap sample and serves as a natural test set for the tree that is fitted to the bootstrap sample. The trees differ because we do not choose the best feasible split for all genes. Instead, we take a sample of a few genes for each node and determine the optimal split on the selected genes. In general, the number of genes selected at each node is the square root of the total number of genes. We assume that the random vector *X* = (*X*_1_, *X*_2_, …, *X*_*k*_) denoting the real-valued predictor variables and the random variable *T* denoting the response variable [38] have an unknown joint distribution *f*_*XT*_ (*X, T*). The main goal is to find a prediction function *f* (*X*) for predicting *T*. The prediction function is determined by minimizing the loss function *L*(*T, f* (*X*)); for classification, the zero-one loss is commonly used.

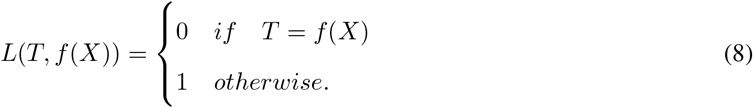

Let *T* = (*x*_1_, *t*_1_), (*x*_2_, *t*_2_), …, (*x*_*N*_, *t*_*N*_) represent the training data set, where *i* = 1, 2, …, *N*. Take a *𝒯*_*m*_ bootstrap sample of size *N* from *𝒯*. Fit a tree using binary recursive partitioning using the bootstrap sample *𝒯*_*m*_ as the training data. Begin by grouping all observations into a single node. Choose *m* predictors randomly from the *p* available predictors, then determine the optimal binary split on the *m* predictors. Finally, using the split, divide the node into two descendant nodes and repeat the process until the stopping criterion is satisfied.

#### C. Statistical Hypothesis on Accuracy

To compare models statistically, the statistical k-fold test was implemented. There were three pair tests considered before implementation. This test is ignored for the Logistic Regression classifier since NBC and Logistic Regression plays the same role. The hypothesis test setup between RF and NBC is:

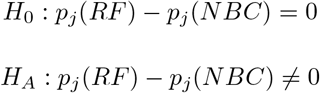

The hypothesis test setup between RF and kNN is:

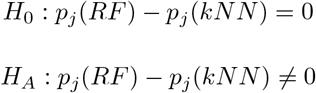

The hypothesis test setup between NBC and kNN is:

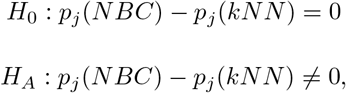

where “*j*” stands for the accuracy. The k-fold cross-validated paired *t* test is applied to compare accuracy. Let’s consider the estimator (e.g., classifiers) for *j* = 1 and *j* = 2. The performance difference among model 1 and 2 in each k-fold cross-validation iteration to get k difference measures are computed. Now, under the null hypothesis that models 1 and 2 perform equally, the following *t* statistic with *k* − 1 degrees of freedom is implemented. This is done in accordance with Student’s *t* test by assuming that these k differences were separately generated and it follows an essentially normal distribution. The test statistic is,

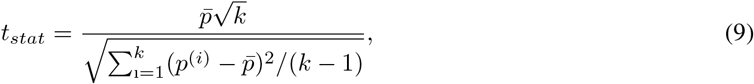

where *p*^(*i*)^ computes the difference between the model performances in the *i*th iteration, *p*^(*i*)^ = *p*_1_^(*i*)^ − *p*_2_^(*i*)^, and 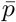 represents the average difference between the classifier performances, 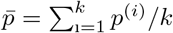. After calculating the *t* statistic, we can compute the *p*-value and compare it to the significance threshold we’ve selected, in this case, 0.05. The null hypothesis will be rejected and acknowledge a substantial difference between the two models if the *p*-value is less than 0.05.

#### D. Model accuracy, 95% Confidence Interval for accuracy, and analysis of Receiver Operating Characteristics curve

The measures for ML model performance are specificity, sensitivity, accuracy, precision, *F*_1_score, and Matthew Correlation Coefficient (MCC). These are described in terms of True Positive as TP, True Negative as TN, False Negative as FN, and False Positive as FP obtained from the confusion matrix. The first measure is,

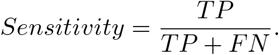

The measures for the sensitivity indicate the likelihood that a diagnostic test will identify persons who truly have the condition. As the sensitivity value rises, the probability of a diagnostic test yielding false-positive results falls. For example, if sensitivity is 95 percent, there is a 95 percent likelihood that the problem will be discovered in this patient. Therefore, utilizing a test with high sensitivity to detect the illness has become standard practice. Next,

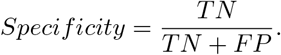

The likelihood that a test would correctly detect a certain condition without producing false-positive results is indicated by the specificity score. If a test’s specificity, for instance, is 95 percent, it means that when we perform a diagnosis for a patient, there is a 95 percent chance that the results will be negative if they don’t have a certain disease condition. Moreover,

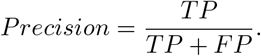

Precision, or the positive predictive value, is the fraction of positive values out of the total predicted positive instances.

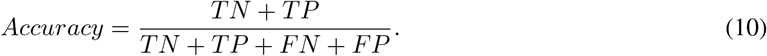

The accuracy score is a measure of the percentage of true positive and true negative outcomes in the chosen population. It’s important to remember that the equation for accuracy indicates that the test’s accuracy may not be as high as its sensitivity and specificity. Accuracy is affected by sensitivity, specificity, and the prevalence of the disease in the target population. A diagnosis may have high sensitivity and specificity but low accuracy for rare illnesses in the population of interest. Moreover, the *F*_1_ score is the harmonic mean of precision and sensitivity; it gives importance to both factors:

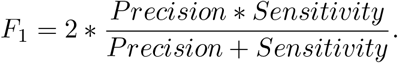

The two measures that are most frequently used in binary classification tasks are accuracy and F1 score, which are computed using confusion matrices. However, especially when applied to unbalanced data sets, these statistical metrics have a serious tendency to provide too optimistic outcomes. A high score is only produced by the Matthews correlation coefficient (MCC) in the event that the prediction was accurate with regard to each of the confusion matrix’s four categories. MCC is defined by,

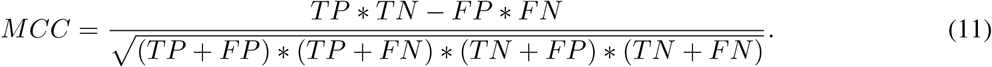

Furthermore, a 95% confidence interval for a machine learning classifier provides a further window into the ambiguity around an ML model’s reported performance, and accuracy [39]. Let’s define the mean test accuracy as *ACC*_*test*_. Therefore, the 100(1 − *α*)% confidence interval for the accuracy of the machine learning model is as follows:

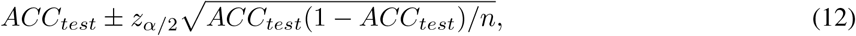

where *n* is the test set size and *z*_*α/*2_ is the critical value obtained for the choice of *α* from the *z*−table.

A direct and natural connection may be made between cost-benefit analysis and diagnostic decision-making using ROC analysis [40]. The Gini Index measures the homogeneity of variables and is related to the ROC. The Gini Index is the area between the ROC curve (AUC) and two times the no-discrimination line (linear). That is, the formula for the Gini Index is: *G*_*i*_ = 2*AUC*–1. Moreover, plotting the true positive rate (TPR) against the false positive rate (FPR) at various threshold levels yields the Receiver Operating Characteristics (ROC) curve [41]. Other names for the true-positive rate include sensitivity, recall, and the likelihood of detection. The probability of a false alarm, which is another name for the false-positive rate, may be computed as (1-specificity). It may also be considered a plot of the power as a function of the decision rule’s Type I Error (estimators of these quantities may be derived from the performance of the population as a whole when that performance is derived from just a sample of the population). After analyzing the ML classification model, the relatively best approach is chosen based on the model accuracy. Mathematically, to describe the behavior of the ROC curve, the following numerical measure is used,

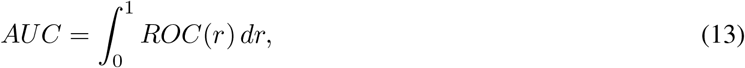

where *r* is the false positive rate. In the next subsection, AI techniques for model explanation are introduced.

#### E. Explainable AI for best predictive model applied to gene expression data

To get the local explanation of the disease with responsible genes, we use Local Interpretable Model-Agnostic Explanations, which is shortly called LIME [23]. How can doctors and patients trust machine learning predictions when each patient is different from the other and multiple parameters can decide between Hepato Cellular Carcinoma or not? To solve this problem, LIME has been used in the test model. It is important that the manner of explanation be relevant to all of the models. Therefore, the researchers have referred to this aspect of the explanation as having a “model-agnostic” status. More precisely, the explanation for a data point **x** is the model *ϕ* that minimizes the locality-aware loss *L*(*f, ϕ*, Π_*x*_), which measures how unfaithful *ϕ* approximates the model to be explained by *f* in its vicinity Π_**x**_, while keeping the model complexity low. Mathematically,

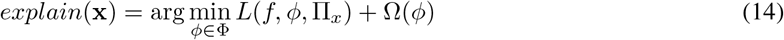

where model *ϕ* belongs to class Φ, and Π_*x*_ is the neighborhood of data point **x**.

Note that models *f* and *ϕ* may operate on different data spaces. The black-box model (function) *f* : *X* → *R* is defined on a large, *p*−dimensional space *X* corresponding to the *p* explanatory variables used in the model. The glass-box model (function) 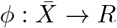 is defined on a *q*−dimensional space 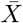 with *q << p*, often called the “space for interpretable representation.” Just like the RF/NBC/kNN models that are trained and fit to the data, the LIME method is used to train this explainer, and then new predictions are made using the *explain*(**x**) function.

## III. Results and Discussion

. After implementing supervised learning on gene expression HCC dataset using RF, NBC, Logistic Regression, and kNN algorithms, the following results were obtained to identify marker genes. Since the dataset was very large, the dimension was reduced using PCA before working on the dataset to keep the important risk factors(genes) only. Then, the dataset was split into training (66.67%) and a testing set (33.33%) by using a simple random sampling technique. The seed was set before splitting for reproducibility. The statistical k-fold test was used to compare models statistically. Prior to implementation of hypothesis testing, three pair-tests were taken into account, and the obtained p-value is less than 0.00001 for each pair of tests, which implies null hypotheses are rejected at the 5% level of significance. Therefore, NBC, RF, and k-NN are unique machine learning models for gene expression liver cancer classification.

In Fig. 3, the most and least essential variables are arranged from top to bottom, with variables with high mean decrease values being relevant. Mean decrease accuracy and Gini Index are used to identify which variables are important. Variance importance ranking in Fig. 3 uses the mean decrease Gini Index to determine which variables (genes in this case) are important. Then a variable importance ranking plot was also plotted. The RF was applied to the data set. Then RF was compared with NBC, Logistic Regression, and k-NN models. Table I shows the performance of the ML algorithms. A ROC curve is a graph that shows how a binary classifier system’s diagnostic capacity changes when its discriminating threshold is altered. Hit rate, which is deviant to true positive or sensitivity, is on the y-axis, and specificity, which is deviant to false positive, is on the x-axis. As a result, ROC might be seen as a power plot that depends on a Type I error. The ROC curves were plotted for RF, NBC, kNN, and Logistic Regression based on the most important predictors determined by PCA for comparing the 1-specificity versus sensitivity.

**Fig. 1:**
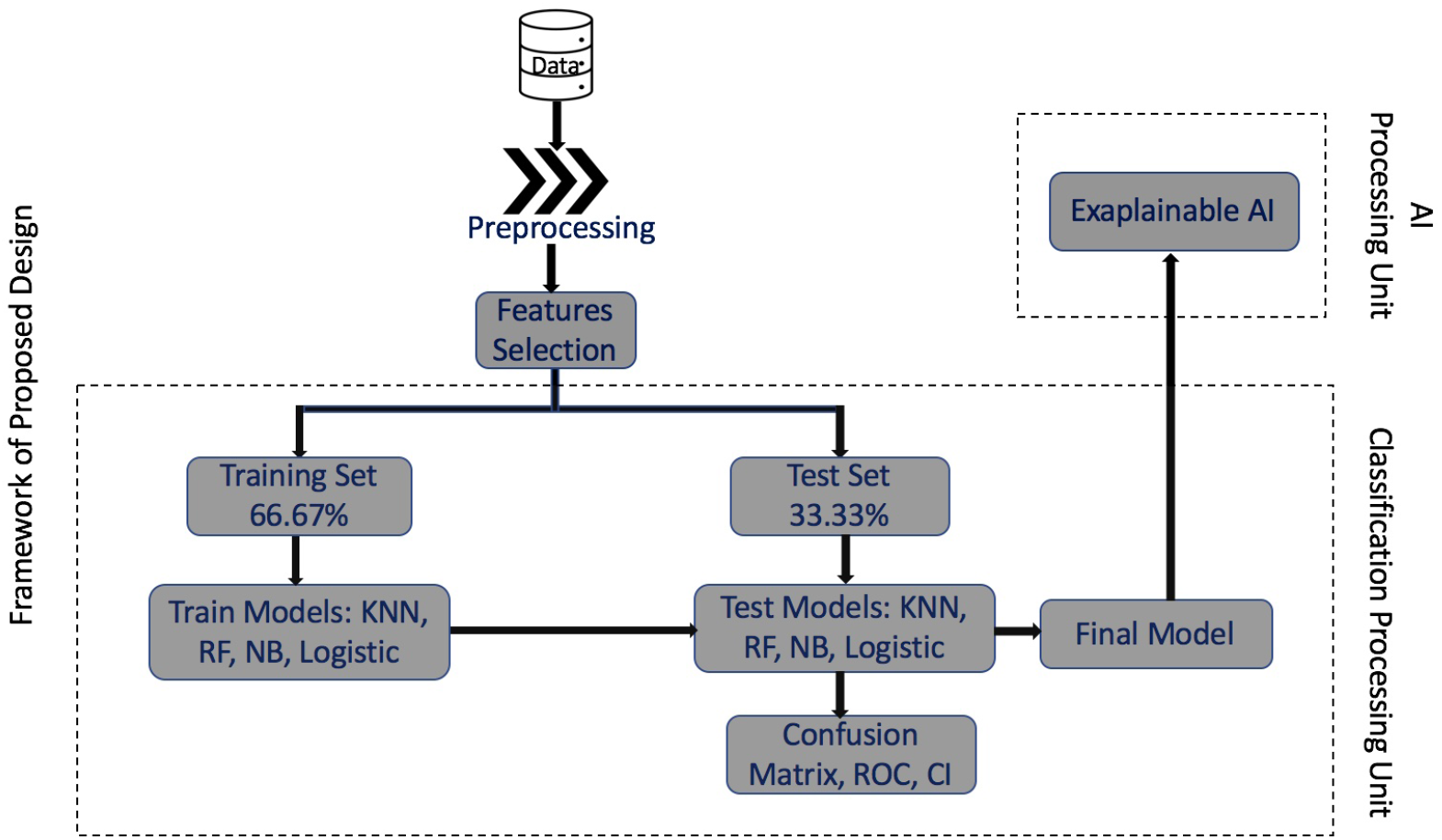
Diagram for ML/AI approaches for microarray analysis of gene expression data. There are two main process steps, first one is classification processing unit where train and test data are used to select the best model, and second one is AI processing unit for global explanation.

**Fig. 2:**
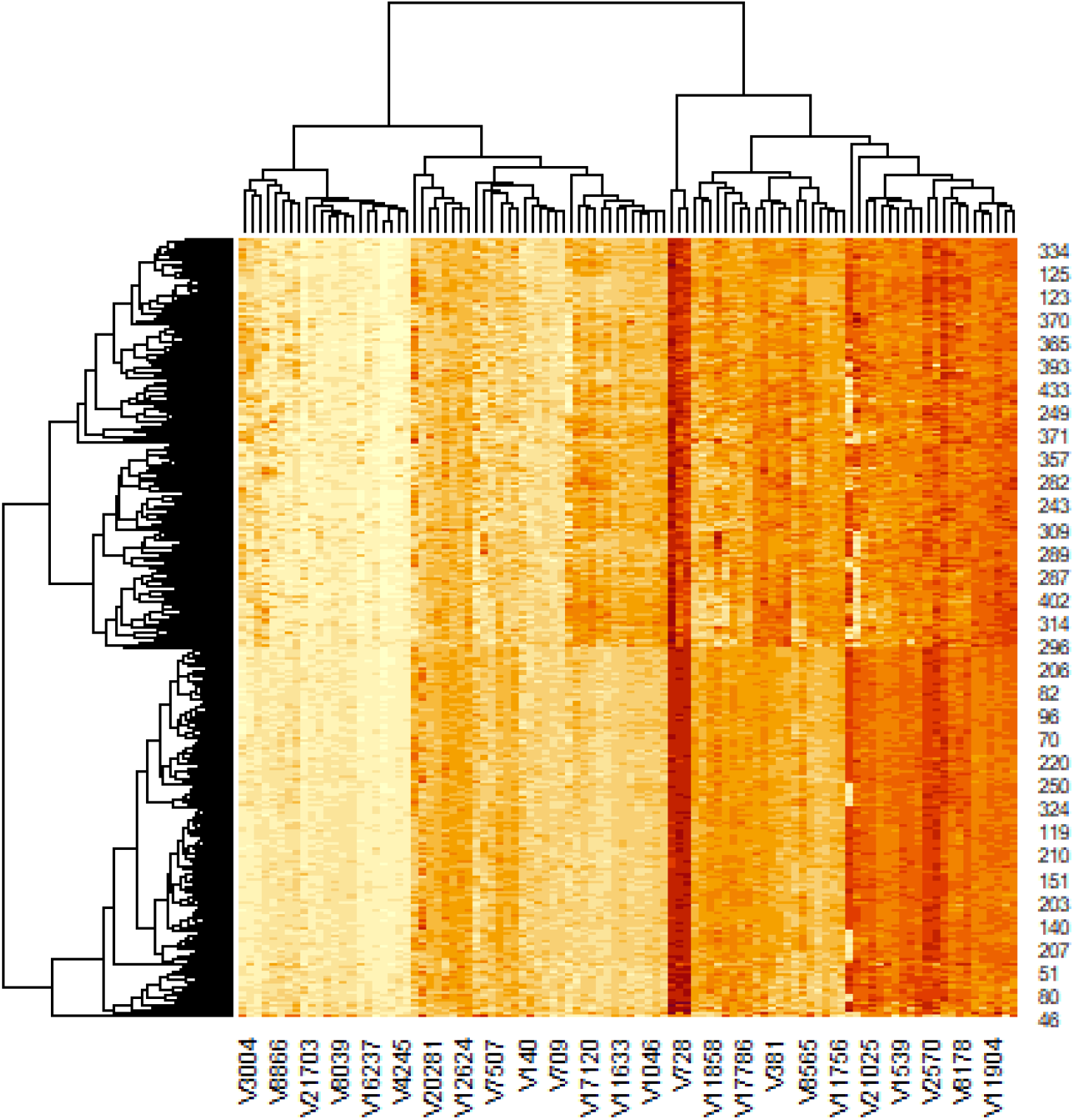
Heatmap of top 100 genes. Due to lack of space this plot represents only 25 gene names in the horizontal line. In the right vertical line, it gives the gene identity numbers from the collection of the data set.

**Fig. 3:**
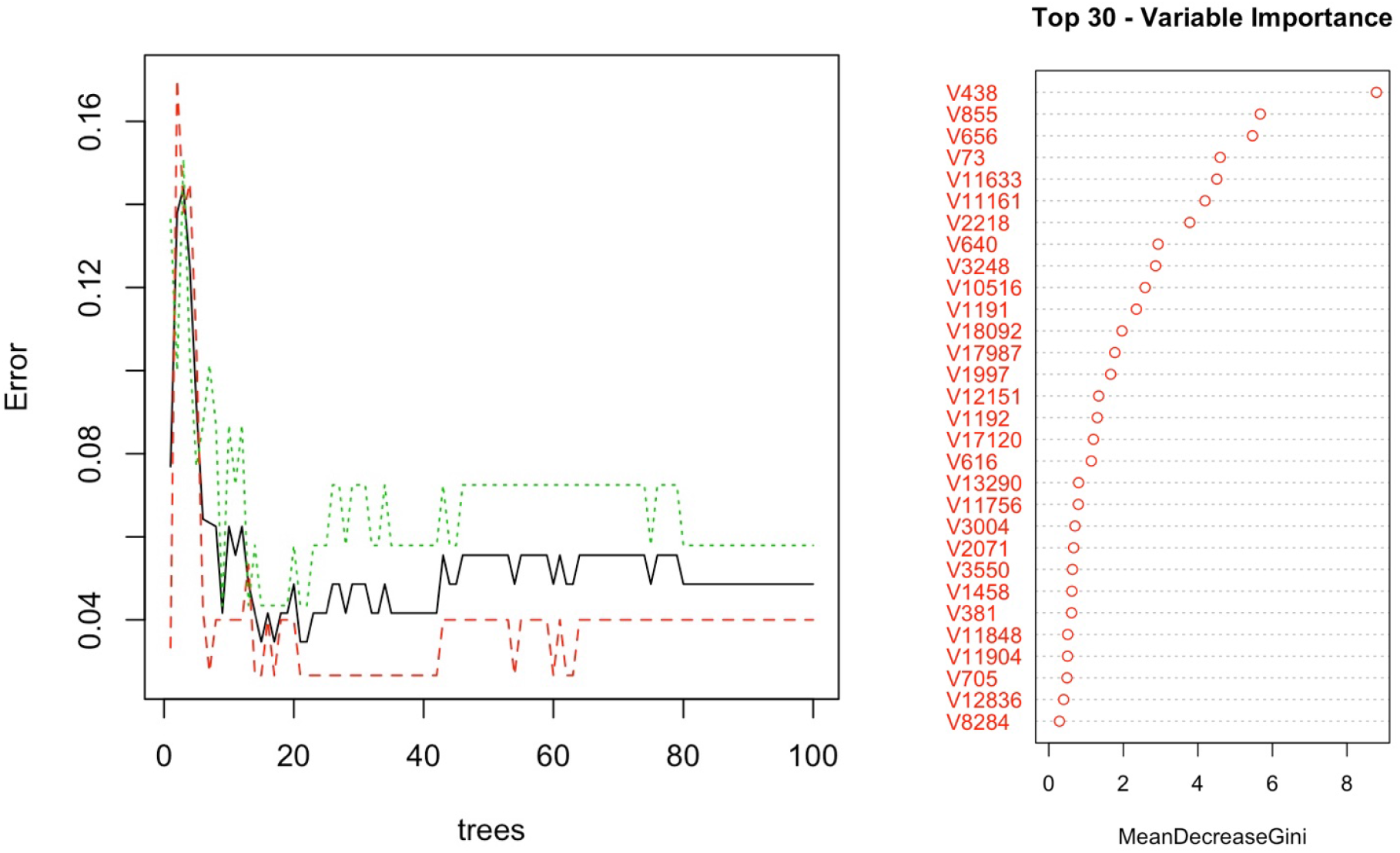
Left panel indicates errors for a growing number of trees in random forest algorithm. For different values of the mtry parameter, the random forest error rates (calculated from out-of-bag situations) are shown as a function of the number of trees. This is based on the random forest voting method used to estimate gene expression response 1 in early testing. The right panel indicates the top 30 variables calculated from the Gini index.

**TABLE I:** Performance for HCC gene expression data classifiers

| Methods | *Sensitivity* | *Specificity* | *Precision* | $F_1$ | $MCC$ |
| --- | --- | --- | --- | --- | --- |
| RF | 0.9600 | 0.9710 | 0.9730 | 0.9664 | 0.9303 |
| NBC | 0.96 | 0.9565 | 0.96 | 0.96 | 0.9165 |
| Logistic Regression | 0.96 | 0.9565 | 0.96 | 0.96 | 0.9165 |
| kNN | 0.9263 | 0.9828 | 0.9888 | 0.9565 | 0.9117 |

Fig. 4. shows the comparison of confusion matrices where the random forest classifier produces the best results. Random Forest has the lowest false positive and false negative cases, and the k-NN classifier has the highest false positive and false negative cases. A histogram of Fig. 5. describes the comparison of models perfectly. The accuracy score of the random forest classifier is very close to 1. Among all, k-NN shows the lowest accuracy, which is 94.92%. Overall, the accuracy level of each classifier performs very well. Table I gives the performances of the ML classifiers using different measures. All measures are very important for explaining the model behaviors and classification performance. An *F*_1_ score is defined as the harmonic mean between precision and recall. It is used as a statistical measure to rate performance. Almost all the models have yielded a 96% *F*_1_ score. RF shows an *F*_1_ score of 0.9664, which means the harmonic mean between precision and recall is 0.9664. Moreover, the MCC scores are present in Table I. The MCC score is above 90%, which suggests all the models produce highly accurate predictions. Fig. 6. shows the diagnostic performance of the applied ML models mentioned above. The AUC under ROC gives almost 0.96, 0.95, and 0.94 for RF, NBC/Logistic Regression, and k-NN, respectively. As diagnostic test accuracy improves, the value gets closer to 1.0, which is the gold standard AUC(AUC=1). Since the sensitivity and specificity are calculated nonparametrically, so the nonparametric approach produces a jagged curve. The metric that effectively averages diagnostic accuracy over the range of test results is summarized by the area under the ROC curve overall. The normal approximation interval is the easiest and most conventional method of generating confidence intervals. We construct the confidence interval using this approach from a single training-test split. Due to the high cost of model training in deep learning, this is particularly appealing. When we want a particular model, it also is interesting when compared to models fit on different training folds, like in k-fold cross-validation, mostly in a deep learning context. The accuracy of the test set shows the expected generalization accuracy. The 95% confidence interval presents us with a measure of the degree of uncertainty surrounding this estimate. Therefore, the 100(1 − *α*)% confidence interval for the accuracy of the machine learning models RF, NBC, Logistic Regression and kNN are calculated. For RF, the 100(1 − *α*)% confidence interval is

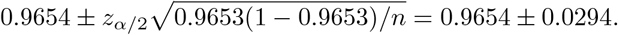

For NBC and Logistic Regression, the 100(1 − *α*)% confidence interval remains the same, which is

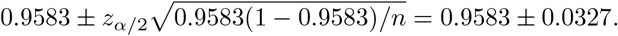

For kNN model, the 100(1 − *α*)% confidence interval is

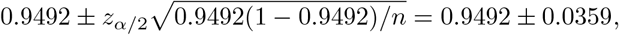

where *n* = 144 is the test set size. *z*_*α/*2_ = 1.96 is the critical value obtained for the choice of *α* = 0.05 from the *z*−table. The sensitivity/specificity pair associated with each point on the ROC curve corresponds to a specific decision threshold. From Fig. 6, the ROC curve that goes through the upper left corner indicates a test that has perfect discrimination, i.e., there is no overlap between the two distributions meaning 100% sensitivity and 100% specificity. Random Forest shows almost 96.5%. NBC and Logistic Regression are close to 95%, and k-NN is 94%, approximately.

**Fig. 4:**
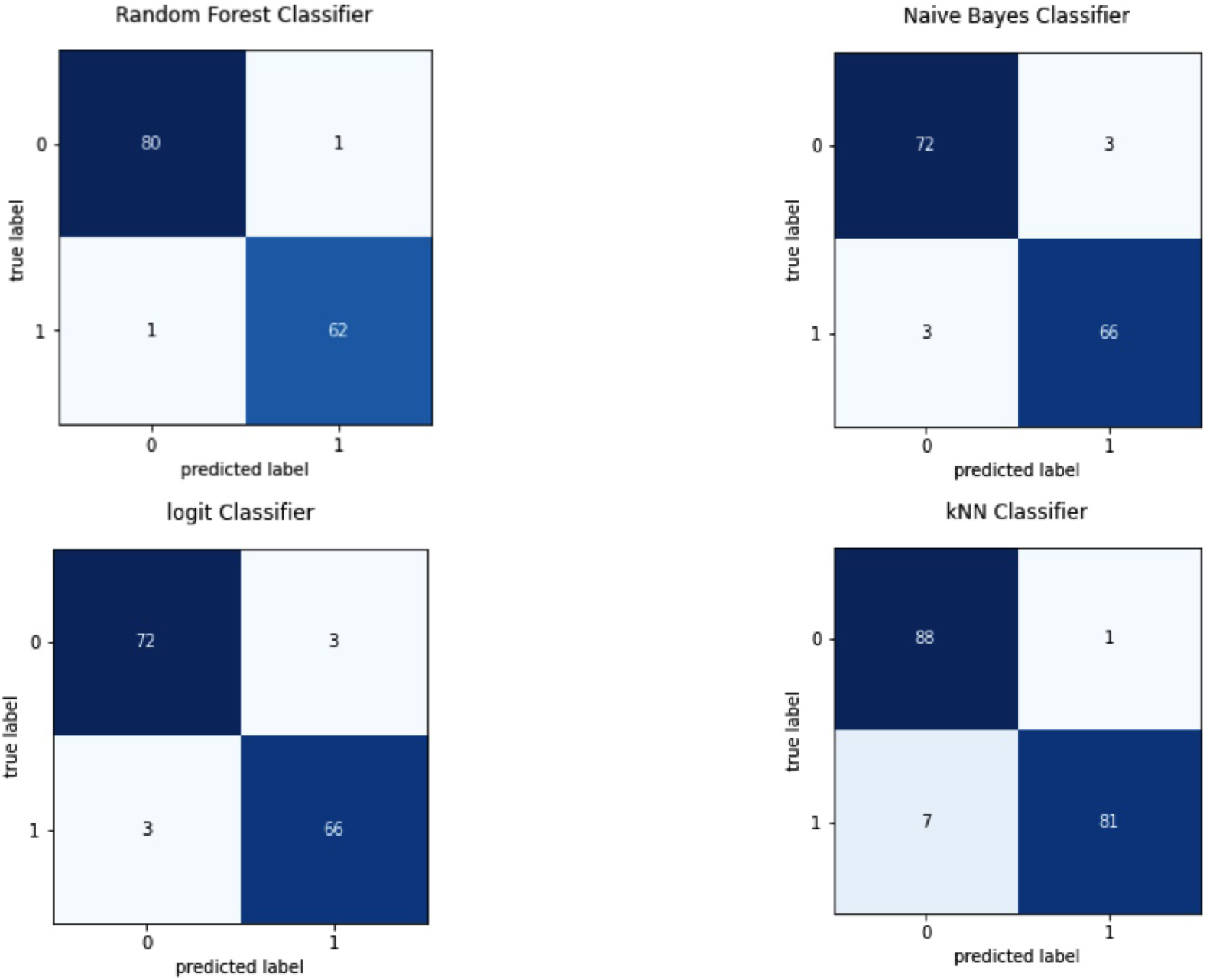
Comparison of Confusion Matrices for four different classifiers. Position (0,0) is the true positive, and (1,1) is the true negative. Position (1,0) is the false positive, and (0,1) is the false negative.

**Fig. 5:**
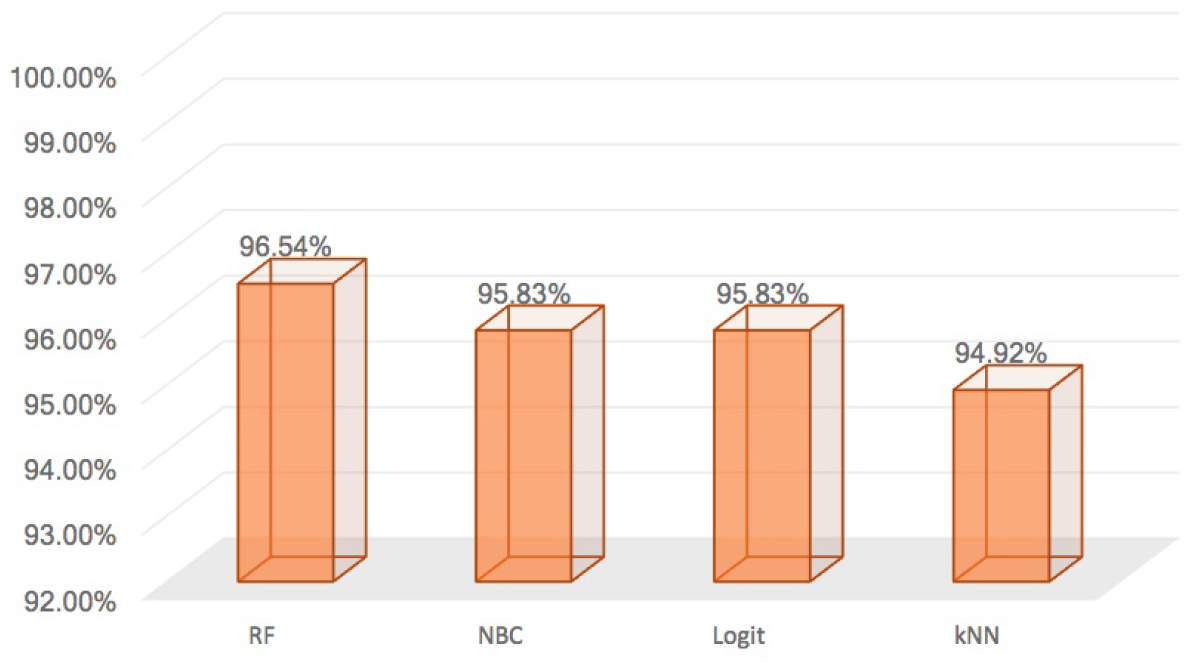
Plot shows the comparison of accuracy for four different classifiers measured in percentage. Names of the ML classifiers are given on the horizontal axis.

**Fig. 6:**
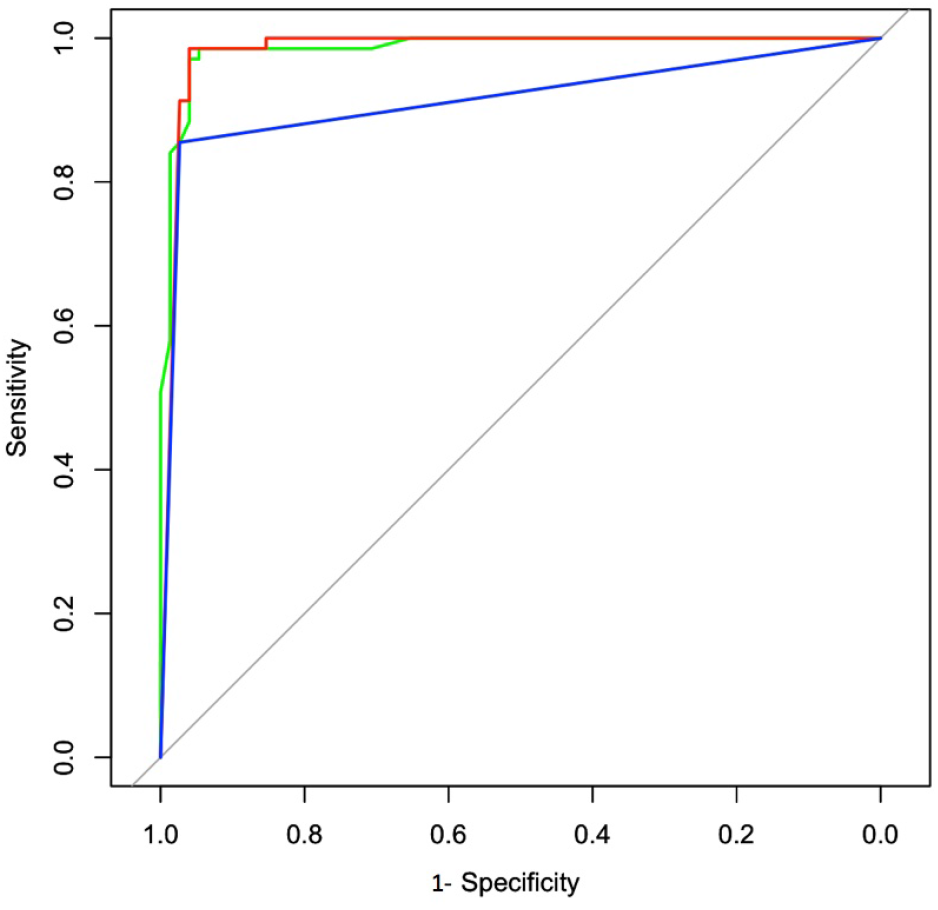
Area under the curve for different ML models. The green line indicates RF, the red line indicates NBC/Logistic Regression, and the blue line indicates kNN classifiers for AUC under ROC curves.

The LIME model focuses on the decision-making process of the machine learning models, hence establishing the base for their use in practical application. The framework analyzes individual observations at the local level. It does not provide a comprehensive explanation for why the model performs as it should but rather explains how well a given observation is classified. The user should be able to comprehend what a model produces if it is interpretable. The responsible genes for HCC are not the same for different patients, so it is essential to know this information for individual patients. As we used tabular data in this research, LIME explains which features influence its decision. The machine learning we used in this research can perform classification based on the attributes of future patients. The patient can be well treated if the physicians know which genes are responsible for HCC. LIME helps us determine the responsible features for HCC and gives us the order of the importance of the features that either positively or negatively impact the response for each particular observation. Fig. 7 depicts two instances of a random forest model predicting that one patient has HCC while the other does not. An “explainer” then explains the prediction by emphasizing the features essential to the model. LIME was used under an ML model with a relatively high accuracy score in this case. After learning about the model’s rationale, the physician is better equipped to decide whether or not to believe it. We chose two patients randomly to apply LIME. Fig. 7 shows the most important 17 features responsible for HCC for those two patients. The model also classified the first person as a positive HCC patient with 98% confidence, and the second person is classified as a negative HCC patient with 98% confidence. For the first patient, out of the first seventeen most important features, fifteen positively impact the response, and two negatively impact the response. For the second patient, thirteen of them are negatively impacting the response, and four of them are positively impacting the response. Since the physicians now know which genes are responsible for HCC for a particular patient, that will help them to treat the patient better.

**Fig. 7:**
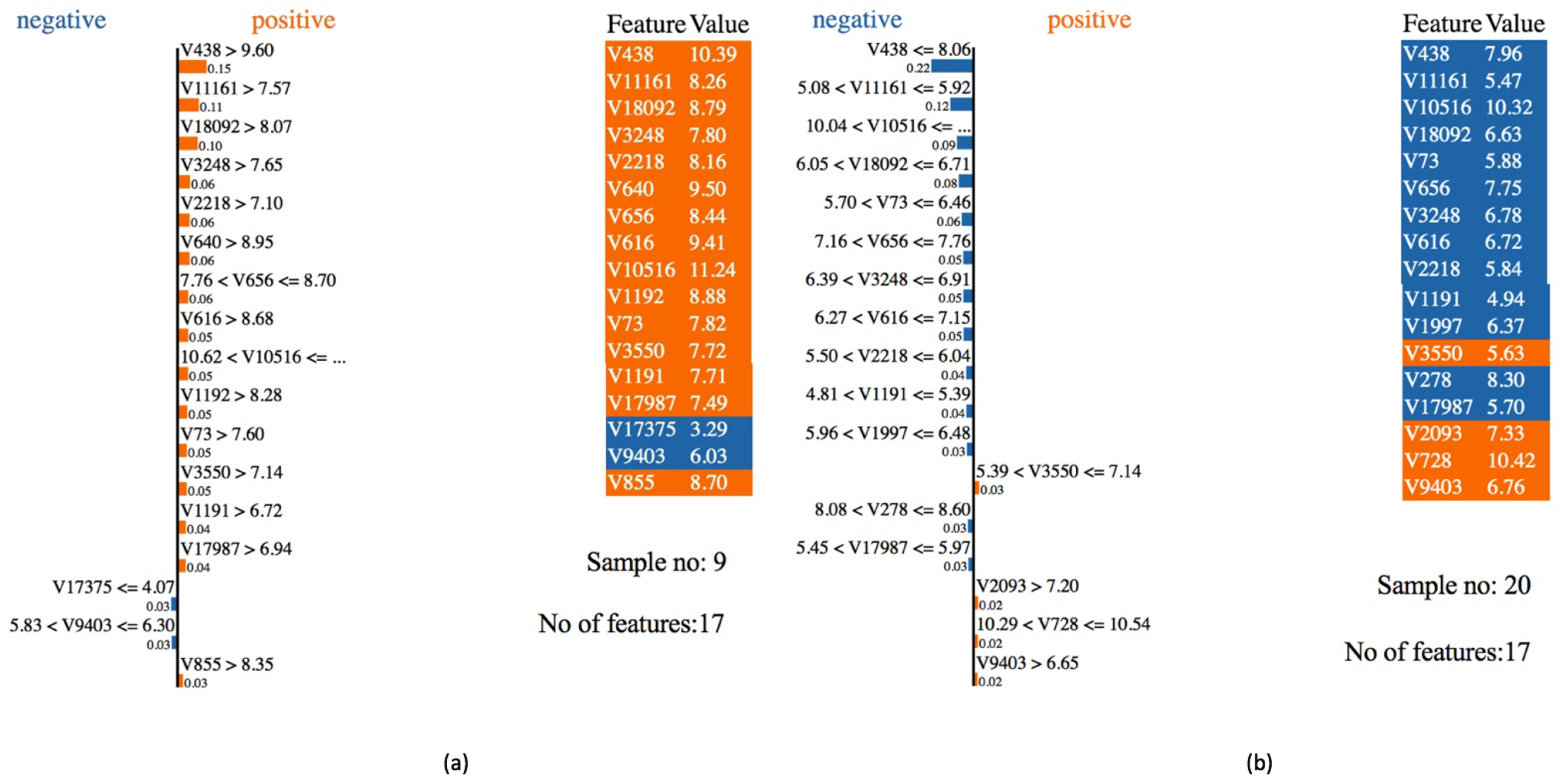
LIME outcomes for the RF classifier for HCC patients. Left panel (a) indicates the credentials of sample-9 of the gene data set. Right panel (b) shows the credentials of sample-20 of the gene data set. The blue color suggests the gene causes negative effects, and the orange color suggests the gene causes positive effects on HCC.

## IV. Conclusions

Our findings supported prior research by proving that machine learning approaches can be utilized to discover responsible genes that have a substantial influence on HCC. According to Lee et al., precision medicine has shown that genetic properties of cancer cells may be used to predict treatment response, and new research suggests that gene-drug links may be predicted very precisely by investigating the cumulative impact of multiple genes at the same time [42]. As a result, the genes responsible for HCC that we discovered using the RF model can assist the development of novel treatments or improve existing therapeutic techniques to prevent HCC in its beginning phases. Because of the limited number of training instances, the availability of a large number of genes, and the various inherent uncertainties, microarray data analysis poses a challenge to conventional machine learning approaches. Since one of the most important advantages of machine learning in the healthcare sector is its capacity to recognize and diagnose illnesses and ailments that would otherwise be challenging to diagnose, this might encompass everything from hereditary diseases to early-stage cancers that are difficult to detect. Due to the high dimensionality of the HCC microarray data, it is necessary to include the features selection to reduce the dimensionality of the data. PCA was used for the features selection, selecting the 100 most essential genes to train the different machine learning models, and using the trained models for the classification using the test data set. The final model was selected with the highest classification accuracy. Based on the model’s classification accuracy, the random forest model was chosen as the final model and fitted the LIME model as the explainable AI model. The explainable AI addresses the challenges of understanding the model at the local level, allowing health professionals to choose whether or not the model should be adopted. When physicians recognize the most critical genes associated with HCC for a particular patient, they can treat patients more effectively. Machine learning models have disadvantages, too, because they overfit most of the time, which may forecast wrong diagnoses. The proposed method may help clinicians to understand the gap between clinical reports and machine intelligent reports based on AI explanation.

## Data Availability

https://www.ncbi.nlm.nih.gov/geo/query/acc.cgi?acc=GSE14520&fbclid=IwAR0iw_BN2d62NEyxHaQVhGBxSXgBcpiZcgipvBYcuzYzpwCTRPPZDG0NDsk

## Institutional Review Board Statement

This study considered a published data set which was publicly available at the https://www.ncbi.nlm.nih.gov [27] to reuse the data set for research purpose and scientific developments.

## Informed Consent Statement

This research was conducted on human subject data. Data were obtained from open sources. The NIH received consent. For more details visit the website [27].

## Data Availability Statement

Data are available online at the https://www.ncbi.nlm.nih.gov/geo/query (accessed on 1 March 2021).

## References

[1] H. B. El-Serag and F. Kanwal, “Epidemiology of hepatocellular carcinoma in the united states: where are we? where do we go?” Hepatology (Baltimore, Md.), vol. 60, no. 5, p. 1767, 2014.

[2] S. Roessler, H.-L. Jia, A. Budhu, M. Forgues, Q.-H. Ye, J.-S. Lee, S. S. Thorgeirsson, Z. Sun, Z.-Y. Tang, L.-X. Qin et al., “A unique metastasis gene signature enables prediction of tumor relapse in early-stage hepatocellular carcinoma patients,” Cancer research, vol. 70, no. 24, pp. 10 202–10 212, 2010.

[3] S. Roessler, E. L. Long, A. Budhu, Y. Chen, X. Zhao, J. Ji, R. Walker, H.-L. Jia, Q.-H. Ye, L.-X. Qin et al., “Integrative genomic identification of genes on 8p associated with hepatocellular carcinoma progression and patient survival,” Gastroenterology, vol. 142, no. 4, pp. 957–966, 2012.

[4] X. Zhao, S. Parpart, A. Takai, S. Roessler, A. Budhu, Z. Yu, M. Blank, Y. E. Zhang, H.-L. Jia, Q.-H. Ye et al., “Integrative genomics identifies yy1ap1 as an oncogenic driver in epcam+ afp+ hepatocellular carcinoma,” Oncogene, vol. 34, no. 39, pp. 5095–5104, 2015.

[5] Y. Wang, B. Gao, P. Y. Tan, Y. A. Handoko, K. Sekar, A. Deivasigamani, V. P. Seshachalam, H.-Y. Ouyang, M. Shi, C. Xie et al., “Genome-wide crispr knockout screens identify ncapg as an essential oncogene for hepatocellular carcinoma tumor growth,” The FASEB Journal, vol. 33, no. 8, pp. 8759–8770, 2019.

[6] Y. Lu, W. Xu, J. Ji, D. Feng, C. Sourbier, Y. Yang, J. Qu, Z. Zeng, C. Wang, X. Chang et al., “Alternative splicing of the cell fate determinant numb in hepatocellular carcinoma,” Hepatology, vol. 62, no. 4, pp. 1122–1131, 2015.

[7] S. Chen, H. Fang, J. Li, J. Shi, J. Zhang, P. Wen, Z. Wang, H. Yang, S. Cao, H. Zhang et al., “Microarray analysis for expression profiles of lncrnas and circrnas in rat liver after brain-dead donor liver transplantation,” BioMed research international, vol. 2019, 2019.

[8] S.-l. Chen, Z.-x. Zhu, X. Yang, L.-l. Liu, Y.-f. He, M.-m. Yang, X.-y. Guan, X. Wang, and J.-p. Yun, “Cleavage and polyadenylation specific factor 1 promotes tumor progression via alternative polyadenylation and splicing in hepatocellular carcinoma,” Frontiers in cell and developmental biology, vol. 9, p. 616835, 2021.

[9] M. Hansen, R. Dubayah, and R. DeFries, “Classification trees: an alternative to traditional land cover classifiers,” International journal of remote sensing, vol. 17, no. 5, pp. 1075–1081, 1996.

[10] C. Huang, L. Davis, and J. Townshend, “An assessment of support vector machines for land cover classification,” International Journal of remote sensing, vol. 23, no. 4, pp. 725–749, 2002.

[11] J. Rogan, J. A. Miller, D. A. Stow, J. Franklin, L. M. Levien, and C. Fischer, “Land-cover change monitoring with classification trees using landsat tm and ancillary data,” Photogrammetric Engineering and Remote Sensing, vol. 69, pp. 793–804, 2003.

[12] G. M. Foody, “Land cover classification by an artificial neural network with ancillary information,” International Journal of Geographical Information Systems, vol. 9, no. 5, pp. 527–542, 1995. [Online]. Available: https://doi.org/10.1080/02693799508902054

[13] M. A. Friedl and C. E. Brodley, “Decision tree classification of land cover from remotely sensed data,” Remote sensing of environment, vol. 61, no. 3, pp. 399–409, 1997.

[14] P. M. Atkinson and A. R. Tatnall, “Introduction neural networks in remote sensing,” International Journal of remote sensing, vol. 18, no. 4, pp. 699–709, 1997.

[15] G. M. Foody and A. Mathur, “A relative evaluation of multiclass image classification by support vector machines,” IEEE Transactions on geoscience and remote sensing, vol. 42, no. 6, pp. 1335–1343, 2004.

[16] L. Breiman and R. Ihaka, Nonlinear discriminant analysis via scaling and ACE. Department of Statistics, University of California Davis One Shields Avenue …, 1984.

[17] L. Breiman, “Randomizing outputs to increase prediction accuracy,” Machine Learning, vol. 40, no. 3, pp. 229–242, 2000.

[18] E. M. Kleinberg, “On the algorithmic implementation of stochastic discrimination,” IEEE Transactions on Pattern Analysis and Machine Intelligence, vol. 22, no. 5, pp. 473–490, 2000.

[19] H. Zhang, “The optimality of naive bayes,” Aa, vol. 1, no. 2, p. 3, 2004.

[20] R. Caruana and A. Niculescu-Mizil, “An empirical comparison of supervised learning algorithms,” in Proceedings of the 23rd international conference on Machine learning, 2006, pp. 161–168.

[21] M. S. Santos, P. H. Abreu, P. J. García-Laencina, A. Simão, and A. Carvalho, “A new cluster-based oversampling method for improving survival prediction of hepatocellular carcinoma patients,” Journal of biomedical informatics, vol. 58, pp. 49–59, 2015.

[22] U. R. Acharya, O. Faust, F. Molinari, S. V. Sree, S. P. Junnarkar, and V. Sudarshan, “Ultrasound-based tissue characterization and classification of fatty liver disease: A screening and diagnostic paradigm,” Knowledge-Based Systems, vol. 75, pp. 66–77, 2015.

[23] M. T. Ribeiro, S. Singh, and C. Guestrin, ““why should i trust you?” explaining the predictions of any classifier,” in Proceedings of the 22nd ACM SIGKDD international conference on knowledge discovery and data mining, 2016, pp. 1135–1144.

[24] I. Palatnik de Sousa, M. Maria Bernardes Rebuzzi Vellasco, and E. Costa da Silva, “Local interpretable model-agnostic explanations for classification of lymph node metastases,” Sensors, vol. 19, no. 13, p. 2969, 2019.

[25] N. B. Kumarakulasinghe, T. Blomberg, J. Liu, A. S. Leao, and P. Papapetrou, “Evaluating local interpretable model-agnostic explanations on clinical machine learning classification models,” in 2020 IEEE 33rd International Symposium on Computer-Based Medical Systems (CBMS). IEEE, 2020, pp. 7–12.

[26] K. Davagdorj, M. Li, and K. H. Ryu, “Local interpretable model-agnostic explanations of predictive models for hypertension,” in Advances in Intelligent Information Hiding and Multimedia Signal Processing. Springer, 2021, pp. 426–433.

[27] W3Techs, “Geo accession viewer,” 2010. [Online]. Available: https://www.ncbi.nlm.nih.gov/geo/query/acc.cgi?acc=GSE14520&amp;fbclid=IwAR0iwBN2d62NEyxHaQVhGBxSXgBcpiZcgipvBYcuzYzpwCTRPPZDG0NDsk

[28] H. Abdi and L. J. Williams, “Principal component analysis,” Wiley interdisciplinary reviews: computational statistics, vol. 2, no. 4, pp. 433–459, 2010.

[29] C. Strobl, A.-L. Boulesteix, T. Kneib, T. Augustin, and A. Zeileis, “Conditional variable importance for random forests,” BMC bioinformatics, vol. 9, no. 1, pp. 1–11, 2008.

[30] L. E. Raileanu and K. Stoffel, “Theoretical comparison between the gini index and information gain criteria,” Annals of Mathematics and Artificial Intelligence, vol. 41, no. 1, pp. 77–93, 2004.

[31] S. Tangirala, “Evaluating the impact of gini index and information gain on classification using decision tree classifier algorithm,” International Journal of Advanced Computer Science and Applications, vol. 11, no. 2, pp. 612–619, 2020.

[32] K. M. Leung, “Naive bayesian classifier,” Polytechnic University Department of Computer Science/Finance and Risk Engineering, vol. 2007, pp. 123–156, 2007.

[33] M. Langarizadeh and F. Moghbeli, “Applying naive bayesian networks to disease prediction: a systematic review,” Acta Informatica Medica, vol. 24, no. 5, p. 364, 2016.

[34] P. Komarek, Logistic regression for data mining and high-dimensional classification. Carnegie Mellon University, 2004.

[35] A. Mucherino, P. J. Papajorgji, and P. M. Pardalos, “K-nearest neighbor classification,” in Data mining in agriculture. Springer, 2009, pp. 83–106.

[36] J. Laaksonen and E. Oja, “Classification with learning k-nearest neighbors,” in Proceedings of international conference on neural networks (ICNN’96), vol. 3. IEEE, 1996, pp. 1480–1483.

[37] L. Jiang, Z. Cai, D. Wang, and S. Jiang, “Survey of improving k-nearest-neighbor for classification,” in Fourth international conference on fuzzy systems and knowledge discovery (FSKD 2007), vol. 1. IEEE, 2007, pp. 679–683.

[38] C. Zhang and Y. Ma, Ensemble machine learning: methods and applications. Springer, 2012.

[39] P. Steinbach, F. Gernhardt, M. Tanveer, S. Schmerler, and S. Starke, “Machine learning state-of-the-art with uncertainties,” arXiv preprint 2204.05173, 2022.

[40] T. Fawcett, “An introduction to roc analysis,” Pattern recognition letters, vol. 27, no. 8, pp. 861–874, 2006.

[41] K. H. Zou, A. J. O’Malley, and L. Mauri, “Receiver-operating characteristic analysis for evaluating diagnostic tests and predictive models,” Circulation, vol. 115, no. 5, pp. 654–657, 2007.

[42] B. K. B. Lee, K. H. Tiong, J. K. Chang, C. S. Liew, Z. A. Abdul Rahman, A. C. Tan, T. F. Khang, and S. C. Cheong, “Design: connecting gene expression with therapeutics for drug repurposing and development,” BMC genomics, vol. 18, no. 1, pp. 1–11, 2017.

